# Evaluating the Effectiveness of Potential Interventions for Guinea Worm Disease in Dogs in Chad Using Simulations

**DOI:** 10.1101/2023.05.22.23290350

**Authors:** Yifan Wang, Tyler Perini, Pinar Keskinocak, Hannah Smalley, Julie Swann, Adam Weiss

## Abstract

Guinea worm disease (or dracunculiasis) is currently transmitted among dogs in Chad, which presents risks for the human population. We studied how interventions implemented at different levels might reduce the spread of Guinea worm disease (geographically and over time) and what levels of interventions might accelerate elimination. We built a multi-water-source agent-based simulation model to analyze the disease transmission among dogs in Chad, as well as in geographic district clusters, and validated it using local infection data. We considered two interventions: (i) tethering, where infected dogs are kept on a leash during periods of infectivity, and (ii) Abate^®^, under which the water source is treated to reduce infectivity. Our results showed that elimination (0 dog infections) is most likely achieved within five years with extremely high levels of tethering (95%) and Abate (90%), when intervention levels are uniform across district clusters. We used an optimization model to determine an improved strategy, with intervention levels which minimize the number of dogs newly infected in the sixth year, under limitations on intervention levels across clusters; the number of dogs infected after five years of intervention could be reduced by approximately 220 dogs with an optimized strategy. Finally, we presented strategies that consider fairness based on intervention resource levels and outcomes. Increased tethering and Abate resources above historical levels are needed to achieve the target of Guinea worm disease elimination; optimization methods can inform how best to target limited resources and reach elimination faster.

## INTRODUCTION

In 2022, only 13 human infection cases with Dracunculiasis (caused by the nematode *Dracunculus medinensis*), more commonly referred to as Guinea worm (GW), were reported in the world; 6 of these cases were in Chad. While human infections are relatively low, 521 dog infections were reported in Chad in the same year.^1^ Dogs are likely playing an indirect role in passing GW to the human population through shared water sources. There have been significant reductions in the numbers of human and dog infections (17 human and 1,040 dog infections reported in Chad in 2018)^2^ thanks in part to two interventions commonly used to reduce the spread of GW disease among dogs, namely Abate treatment of water sources and tethering of dogs.^1^ In this study, we developed a simulation model and assessed how different levels of these interventions across multiple geographic areas might further reduce the spread of GW disease.

Computer simulation models have been used for modeling the epidemiology of influenza,^3,4^ HIV/AIDS,^5^ and malaria.^6,7^ Perini et al.^8^ presented a stochastic agent-based simulation model capturing the natural transmission pathway(s) of GW infections in dogs in Chad. The model accounted for environment-driven seasonality, an important factor in GW transmission^9,10^ and was validated using integrated national data provided by the Chad Guinea Worm Eradication Program (GWEP). Other models on GW infections are predominantly deterministic compartmental models which are not validated using empirical data^11-13^ or consider human hosts only. ^14-16^

It is important that interventions to reduce the prevalence of GW disease in dogs in Chad are informed by the dynamics of GW transmission, with the goal of ultimately eliminating the disease.^11^ Two interventions commonly used for reducing GW infections are tethering of dogs and Abate treatment of water sources. When an infected dog is tethered, it is kept on a leash for at least 30 days after the emergence of a worm to prevent this dog from infecting any water sources. Abate is the use of organophosophate larvicide temephos (ABATE Larvicide, BASF, or just “Abate”) to kill the infectious copepods in the water which dogs consume via drinking the water and eating paratenic and/or transport hosts such as fish and frogs.^17^ Once applied, Abate is able to suppress copepod populations for up to 28 days.

The number of GW infection cases per 1,000 dog population per year differ among regions in Chad and regional infection data suggests that intervention coverages are not uniform across the country.^18^ Moreover, dogs in different regions may face different levels of infection risk. For example, an ecological study on dogs in Chad measured significant regional differences in access to ponds, diets, and average range of travel from households.^19^ Thus, it is important to design an intervention approach that accounts for multiple water sources (MWS) or regions. The model presented by Perini et al.^8^ considered a single shared water source among all dogs, which is likely to overestimate infections due to inherent homogeneous mixing assumptions. We developed an MWS agent-based simulation model incorporating environment-driven seasonality (similar to Perini et al.) along with heterogeneous mixing across water sources.^8^

On the ground, the hydrological landscape of Chad is quite complex and includes a rainy and dry season; the former experiences flooding of the Chari River, and as the flooding recedes in the latter, many, smaller water sources (e.g., ponds) are left behind until they evaporate. To capture the geographic diversity while keeping model complexity (and run times) in check, we included three geographic district clusters in Chad, with limited connectivity between clusters. In an extensive computational study, we examined strategies for determining tethering and Abate levels among these clusters, to estimate the percentage of the dog population infected (geographically and over time) under various levels of interventions. When intervention resources are uniform across regions (i.e., equal levels of tethering and Abate), we identified the level of intervention coverages needed to reach nationwide elimination. With the goal of increasing intervention effectiveness, we also developed a simulation optimization approach which suggests region-based intervention levels, considering limited resources for managing the interventions across all regions as well as “fairness” of resource allocation strategies. The results suggest that an optimization-based strategy for interventions across regions might lead to 220 fewer infections after 5 years compared to a uniform strategy. These results highlight the potential benefits of data-driven analytics-based decision making in choosing intervention levels to achieve elimination.

## DATA AND METHODS

### Data

From the Chad GWEP, we obtained information about the number of worms emerging from dogs per month and the number of dogs with an emerging worm per month for the years 2016-2018 within 1674 village, 88 zones, 19 districts and 5 regions of Chad.^18^ We considered “district” as the smallest units for clustering to balance the level of complexity and data quality (of GW infections in each unit).

We clustered districts based on latitude, longitude, position along the Chari River, elevation of each district center, and relative worm emergence per month. For each cluster, we calculated the total number of worms emerging from dogs per month and the number of dogs with an emerging worm per month for the years 2016-2018, then we calibrated infectivity parameters using integrated data from the corresponding cluster. We used the cross-entropy method^20^ to find the best infectivity parameter values to closely match simulation results with actual infection data (i.e., parameters which minimize the mean squared error with respect to dogs exuding and worms exuded). Details of the district clustering procedure and parameter calibration are included in the Supplemental Materials Section 1.

### Agent-Based Multi-Water-Source Simulation Model

The simulation model presented in Perini et al.^8^ simulates the life cycle of GW and daily interactions between the dogs, worms, and water source over multiple years. With some probability, dogs acquire infection from the water source via consumption of water containing copepods which carry the GW larvae, or by consuming (short-term) transport/paratenic hosts, and dogs with patent infections (emergent worms) infect the water source. Each worm released into a water source releases tens of thousands of first-stage GW larvae. The simulation keeps track of the emerging worms and corresponding GW larvae cohorts released into the water. As larvae mature in 10-14 days, the water source becomes more infective. When the larvae cohort begins to die after 30 days, the water source becomes less infective. The probability of a dog acquiring an infection from the water source on a given day, or water infectivity, depends on the number of infective GW larvae in the water, the monthly weather/seasonality factor, and the Abate level.

The probability of a dog accessing a water source, whether to infect or acquire infection, depends on the tethering coverage parameter. Dogs remain susceptible to new infections, whether they are already infected or not, and one or more worms may emerge from an infected dog. In the simulation, (1) each infected dog exudes a number of worms based on the worms per dog distribution, (2) each worm is assigned a worm-emergence day based on the incubation period of 10-14 months, (3) if a dog is re-infected again before the worms emerge, then new/additional worms are created. The simulation tracks the number of dogs infected as well as total worms.

The multi-water-source (MWS) agent-based simulation model we developed builds upon the single water source framework of Perini et al.^8^, with the similar dynamics of GW transmission in dogs, but considering multiple connecting water sources (clusters), where each water source/cluster in the simulation might correspond to multiple smaller bodies of water (e.g., ponds/puddles) in geographic proximity of each other. Each water source/cluster has distinct infectivity parameters, dog populations, and annual intervention coverages (tethering and Abate) from 2016 to 2018. In the model, the traveling behavior of dogs informs the level of interactions between clusters, based on an ecological study of GW infection in dogs.^19^ The pseudocode for the simulation model is presented in the Supplemental Materials Section 2.

### Intervention Methods

Consistent with the current practice^1^ and previous work,^8^ we examined two intervention methods (i.e., tethering and Abate) to evaluate the effectiveness with respect to reducing GW infections in dogs. In the simulation, dogs are tethered with some probability, and tethered dogs do not acquire new infections during this time. Abate treatment is effective for up to 28 days after application. In the simulation, a 20% Abate level, for example, means 20% reduction (on average) in infectivity in the water source. When a dog visits a water source with 20% Abate, the dog has a 20% lower chance (on average) of being infected compared to visiting a water source that has not been treated with Abate.

The main outcome measure in the simulation is the number (or percentage) of dogs (newly) infected annually geographically (across three clusters) and at the end of the intervention horizon (5 years). We analyzed the impact of multiple strategies, evaluated by the main outcome measure, and considering two other factors, namely, *allocation fairness* and *outcome fairness*, which try to minimize the differences between intervention levels (i.e., resources allocated to manage the interventions) or outcome measures across clusters, respectively. The strategies included: (i) *Uniform* strategy, where tethering and Abate levels are the same in each cluster, (ii) Varying levels of interventions in different clusters, and (iii) “Optimized” levels of interventions in each cluster (with side constraints to achieve different levels of allocation or outcome fairness). Note that the uniform strategy maximizes allocation fairness but does not ensure outcome fairness.

Across all scenarios, tethering coverages varied from 45% to 95% (the national average from 2016 to 2018 was 71%; see Supplemental Materials Section 1, Table S1) and Abate levels varied from 15% to 75% (the national average from 2016 to 2018 was 21%). Tethering and Abate levels beyond 95% and 75%, respectively, were deemed as impractical (within this 5 year period) by implementing partners on the ground. However, we also investigated the potential reduction in infections if higher levels of Abate were possible.

### Optimized Strategies

To search for optimized strategies, we adopted the cross-entropy (CE) method presented in Wang et al.^20^ considering different tethering and Abate levels (on average, across multiple clusters) or performance constraints.^21^ The primary objective was to find cluster-specific levels of Abate and tethering to minimize the percentage of dogs infected at the end of the fifth year.

We explored strategies for allocating resources to clusters, assuming resource capacities enabling 70% tethering and 20% Abate levels across all clusters combined but with higher cluster-specific intervention level upperbounds. We also evaluated strategies which maximize allocation fairness and outcome fairness. Additional details are presented in Supplemental Materials Section 3, and the optimization model details are included in the Supplemental Materials Section 4.

## RESULTS

### Clustering and Parameter Calibration

The map in Figure 1 shows the districts identified in the data provided from GWEP which we grouped into 3 clusters (West, East, and Central) as outlined in Table 1, capturing 76.4% of the susceptible dog population and 83.3% of the infected dog population. The East cluster has approximately twice the population of dogs as that of the other clusters we study. The Central cluster is neighboring both the East and West clusters, so dogs may go back and forth between those pairs. See the Supplemental Materials Section 1 for more details.

**Figure 1.**
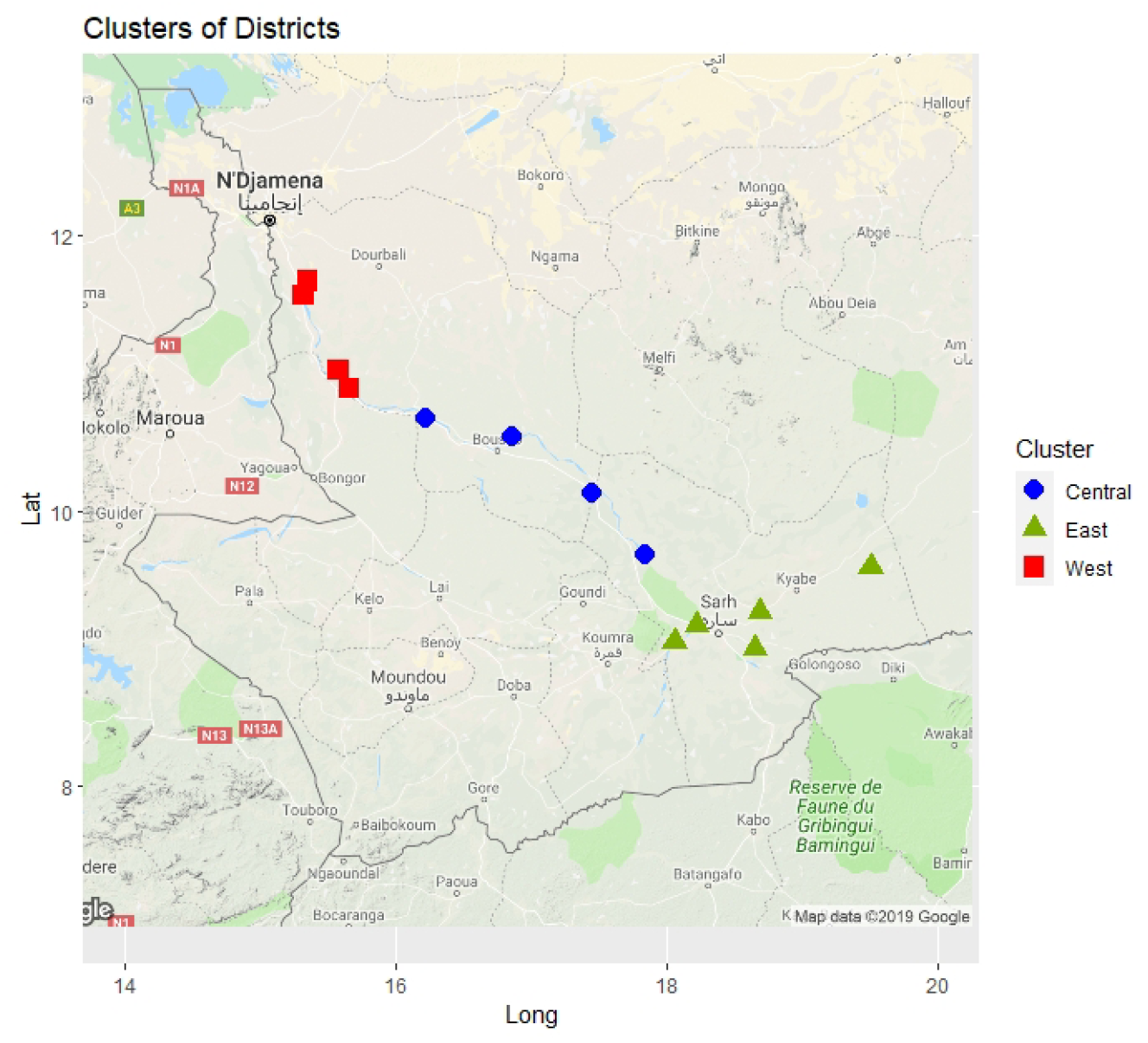
Districts in Chad by computed clusters.

**Table 1.** District clustering result for Chad, with susceptible and infected dog populations from year 2016 to year 2018.

| Cluster | Districts | Susceptible Population<br>(Number of Dogs) | Reported Dogs Infected |  |  |
| --- | --- | --- | --- | --- | --- |
|  |  |  | 2016 | 2017 | 2018 |
| West | Dourbali, Guelendeng,<br>Mandelia, Massenya,<br>Ndjamena-Sud | 8,223 (14.3%) | 261 | 266 | 378 |
| Central | Bailli, Bousso, Korbol,<br>Kouno | 11,910 (20.7%) | 174 | 147 | 272 |
| East | Banda, Bedaya, Biobe,<br>Danamadji, Haraze,<br>Kyabe, Sarh | 23,855 (41.4%) | 512 | 375 | 371 |

The infectivity curves for each of the three clusters are shown in Figure 2. The calibrated disease parameters for each cluster are included in the Supplemental Materials Section 1; the traveling behavior of dogs is reported in Supplemental Materials Section 4. Note that for the East cluster, the number of dogs infected in year 2018 (1.55 % of dogs) places the infectivity rate for the cluster towards the lower end of the infectivity curve, while in both the Central and West clusters the number of dogs infected in year 2018 (2.28% and 4.60%, respectively) places the infectivity rate for these clusters on the higher end of the infectivity curves.

**Figure 2.**
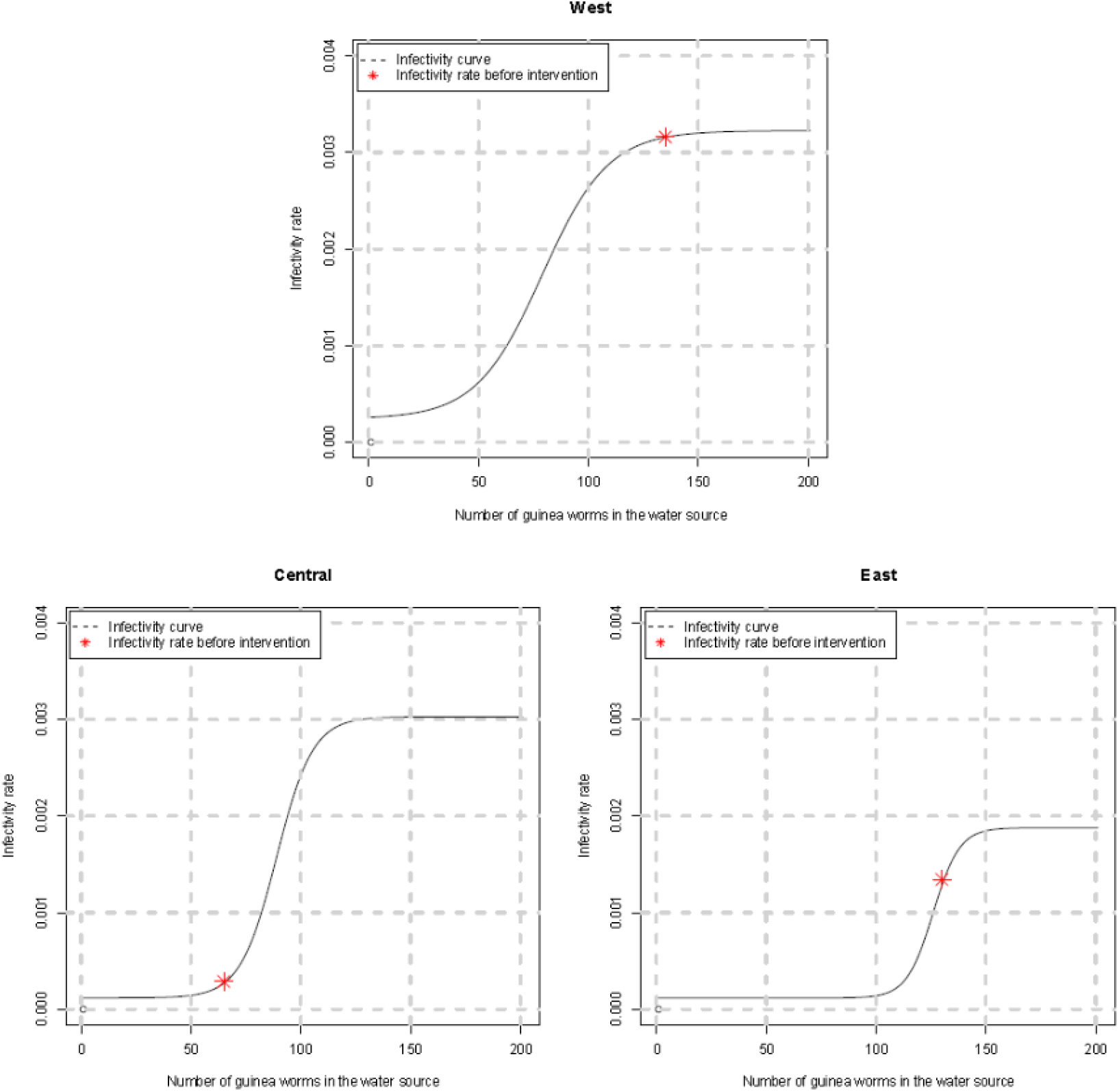
Infectivity curves for the West, Central, and East clusters.

We also included a status quo of the number of GWs in the fourth year (calculated as the maximum worms exuded across all months in 2019), before additional interventions; this is indicated by the asterisks (*) in Figure 2. Note that due to the characteristics of the Central cluster, the cluster’s infectivity rate is towards the lower end of its infectivity curve (e.g., which could be a function of the number/size of water sources, or the behavior of dogs in that cluster). In contrast, the East and West clusters are currently on the middle to upper end of their infectivity curves. In addition, the maximum infectivity increases as you move from East to West, which is associated with decreasing elevation and the direction that the river flows.

Figure 3 shows the observed and simulated number of dogs exuding worms by month over three years for each cluster. Additional details regarding the simulation model are included in the Supplemental Materials Section 2.

**Figure 3.**
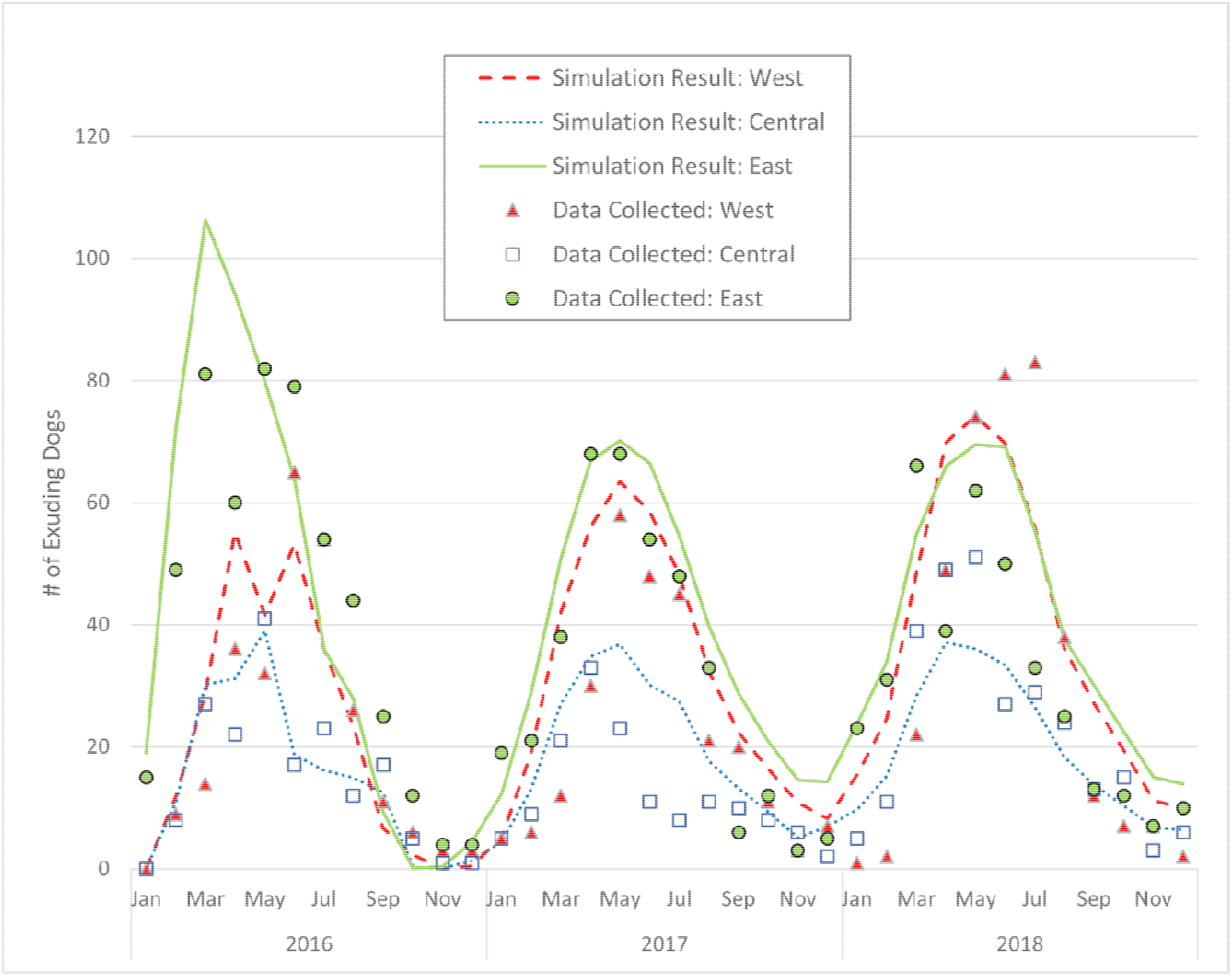
Observed number of dogs infected by cluster for years 2016-2018 and simulated dogs infected from the MWS model.

### Simulated Outcomes under Various Strategies

All test cases and simulation results (mean of 20 replications) are listed in Tables 2 to 4.

**Table 2.** Percentage of dogs infected after uniform interventions for 5 years. Current levels of tethering and Abate (on average) are indicated.

|  |  | Abate Levels (Uniform) |  |  |  |  |  |  |  |  |  |  |  |  |  |  |  |  |
| --- | --- | --- | --- | --- | --- | --- | --- | --- | --- | --- | --- | --- | --- | --- | --- | --- | --- | --- |
|  |  | 15% | 20% | 25% | 30% | 35% | 40% | 45% | 50% | 55% | 60% | 65% | 70% | 75% | 80% | 85% | 90% | 95% |
| Tethering Levels (Uniform) | 45% | 19.76% | 18.09% | 16.75% | 15.30% | 13.14% | 7.35% | 3.91% | 2.45% | 1.52% | 1.27% | 1.12% | 0.95% | 0.80% | 0.61% | 0.44% | 0.28% | 0.10% |
|  | 50% | 18.33% | 16.73% | 14.03% | 11.13% | 6.81% | 4.21% | 2.09% | 1.68% | 1.45% | 1.28% | 1.12% | 0.96% | 0.77% | 0.59% | 0.44% | 0.27% | 0.09% |
|  | 55% | 15.39% | 12.25% | 8.51% | 5.08% | 2.56% | 2.07% | 1.85% | 1.62% | 1.46% | 1.28% | 1.09% | 0.93% | 0.76% | 0.60% | 0.43% | 0.25% | 0.09% |
|  | 60% | 8.70% | 5.85% | 3.34% | 2.53% | 2.22% | 1.96% | 1.76% | 1.59% | 1.41% | 1.24% | 1.11% | 0.93% | 0.75% | 0.59% | 0.42% | 0.25% | 0.07% |
|  | 65% | 3.85% | 2.84% | 2.48% | 2.30% | 2.10% | 1.92% | 1.75% | 1.58% | 1.42% | 1.23% | 1.09% | 0.91% | 0.75% | 0.56% | 0.40% | 0.23% | 0.06% |
|  | 70% | 2.85% | 2.61% | 2.44% | 2.23% | 2.05% | 1.90% | 1.74% | 1.55% | 1.40% | 1.21% | 1.06% | 0.90% | 0.74% | 0.56% | 0.38% | 0.22% | 0.04% |
|  | 75% | 2.73% | 2.56% | 2.35% | 2.17% | 2.03% | 1.86% | 1.71% | 1.55% | 1.39% | 1.23% | 1.06% | 0.87% | 0.70% | 0.53% | 0.36% | 0.18% | 0.02% |
|  | 80% | 2.66% | 2.51% | 2.33% | 2.14% | 2.05% | 1.86% | 1.68% | 1.54% | 1.37% | 1.18% | 1.02% | 0.84% | 0.67% | 0.50% | 0.31% | 0.12% | 0.02% |
|  | 85% | 2.59% | 2.45% | 2.25% | 2.13% | 1.99% | 1.79% | 1.63% | 1.45% | 1.31% | 1.11% | 0.95% | 0.80% | 0.59% | 0.41% | 0.25% | 0.09% | 0.01% |
|  | 90% | 2.54% | 2.35% | 2.20% | 2.06% | 1.87% | 1.68% | 1.53% | 1.34% | 1.18% | 0.97% | 0.82% | 0.65% | 0.47% | 0.26% | 0.14% | 0.03% | 0.003% |
|  | 95% | 2.21% | 2.02% | 1.89% | 1.67% | 1.48% | 1.27% | 1.11% | 0.89% | 0.74% | 0.63% | 0.42% | 0.26% | 0.16% | 0.07% | 0.02% | 0.003% | 0.000% |

**Table 3.**
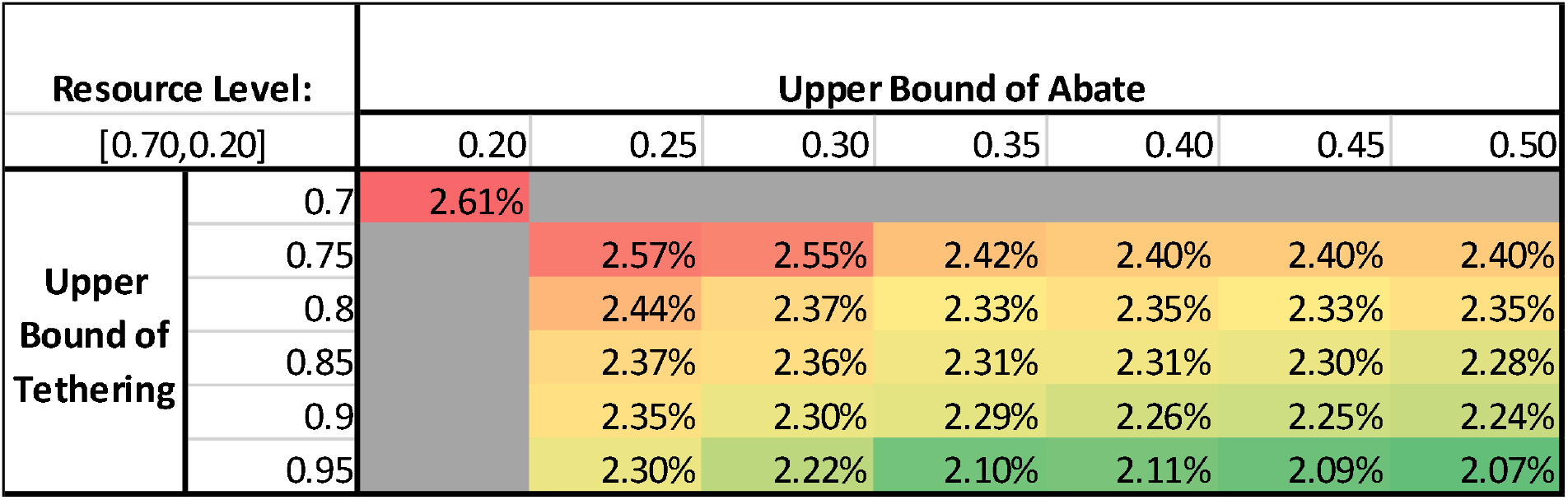
Percentage of dogs infected after using the optimized strategy under different cluster-specific upper bounds for 5 years, with average levels of 70% tethering and 20% Abate across clusters.

| Resource Level: |  | Upper Bound of Abate |  |  |  |  |  |  |
| --- | --- | --- | --- | --- | --- | --- | --- | --- |
| [0.70, 0.20] |  | 0.20 | 0.25 | 0.30 | 0.35 | 0.40 | 0.45 | 0.50 |
| Upper Bound of Tethering | 0.7 | 2.61% |  |  |  |  |  |  |
|  | 0.75 |  | 2.57% | 2.55% | 2.42% | 2.40% | 2.40% | 2.40% |
|  | 0.8 |  | 2.44% | 2.37% | 2.33% | 2.35% | 2.33% | 2.35% |
|  | 0.85 |  | 2.37% | 2.36% | 2.31% | 2.31% | 2.30% | 2.28% |
|  | 0.9 |  | 2.35% | 2.30% | 2.29% | 2.26% | 2.25% | 2.24% |
|  | 0.95 |  | 2.30% | 2.22% | 2.10% | 2.11% | 2.09% | 2.07% |

**Table 4.**
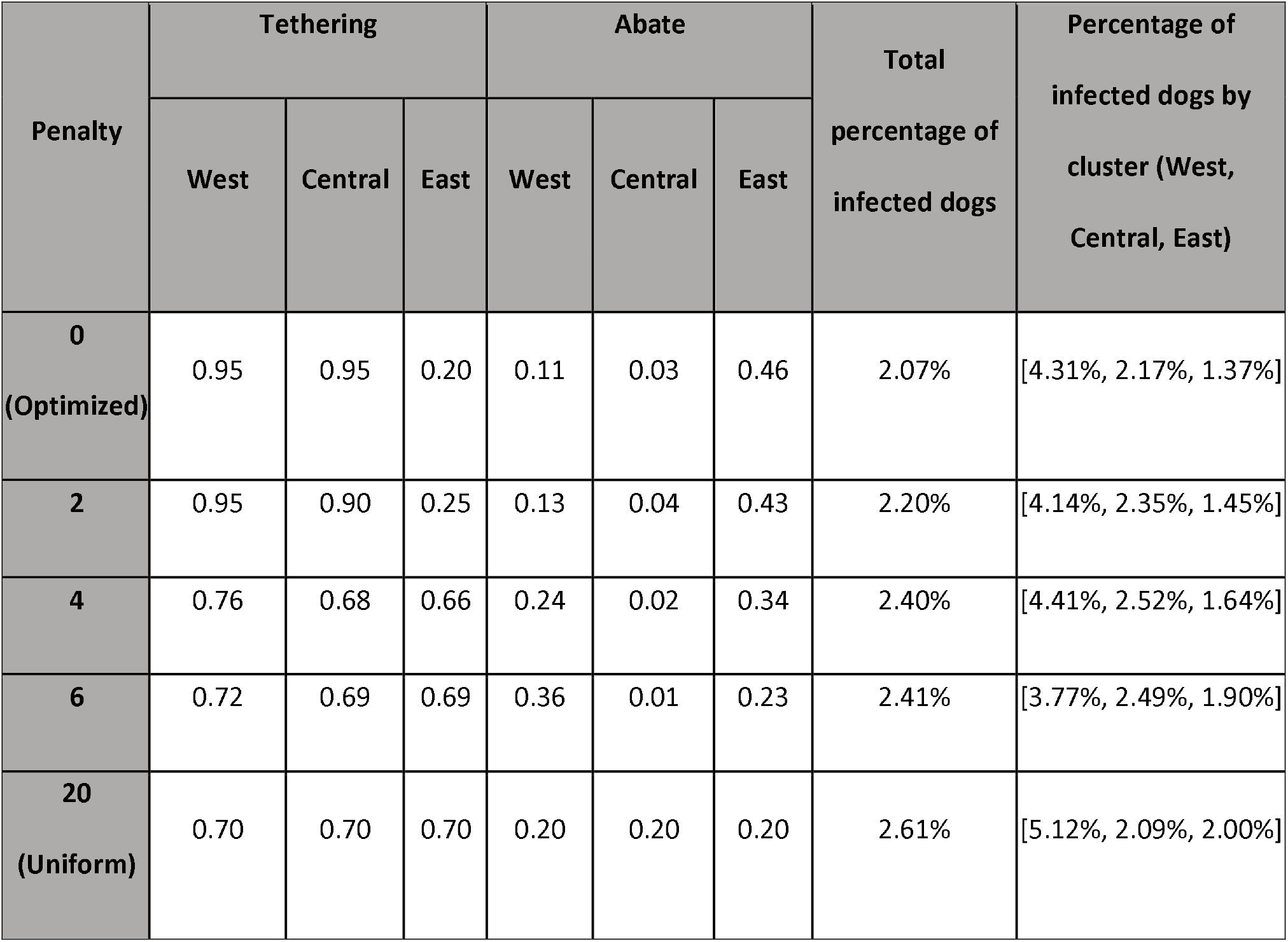
Optimized strategies under different penalty weights for allocation fairness, with the resulting total percentage of infected dogs after five years of intervention both overall and by cluster.

#### Uniform Interventions

In Table 2, we report the percentage of dogs infected at the end of five years for combinations of tethering and Abate that are uniform across all clusters. When tethering and Abate are low (45% and 15%, respectively), the infected dogs total 19.76% of the population, while when tethering and abate are at the high values of 95% and 75%, there are approximately 0.16% dogs infected. 0 dog infections can be achieved with 5 years of interventions but only at higher Abate levels (95% or higher). Results for shorter and longer intervention time periods are reported in Supplemental Materials Section 6.

If tethering coverage continued at 95% and Abate coverage at 75%, the percentage of dogs infected after 9 years of intervention would drop to 0.006%, which is equivalent to 3 dogs. With up to nine years of interventions, 0 dog infections can only be achieved with at least 90% abate levels, unless infected dogs are tethered at levels above 95%. If tethering and Abate are continued at their current levels of approximately 70% and 20% applied uniformly, then the simulation results showed that 2.61% of dogs would be infected after five years of intervention (or 2.57% after nine years). The percentage of infected dogs by cluster for low levels of tethering and Abate (45% and 15% respectively) is relatively low in the East cluster (3.60%), but nearly 40% in the West and Central clusters. High levels of tethering and Abate (85% and 35%, respectively) reduce these percentages to 1.99% overall and 3.56%, 1.61%, and 1.64% for the West, Central, and East clusters, respectively. Notably, the percentage of dogs infected per cluster is much more balanced (i.e., higher levels of outcome fairness) at these higher intervention levels. As seen in Table 2, increasing the level of Abate versus tethering by the same percentage leads to a larger percentage decrease in dog infections.

#### Optimized Intervention Levels

Table 3 shows the percentage of dogs infected after five years of intervention where the intervention levels may differ by cluster. Recall that uniformly applying 70% and 20% tethering and Abate, respectively, results in 2.61% of dogs infected after five years of intervention. This corresponds to 5.12%, 2.09%, and 2.00% infected dogs in the West, Central, and East clusters, respectively. However, these percentages can be reduced by allowing higher levels (upper bounds) of tethering and/or Abate in each cluster, while keeping the “average” levels across the clusters still at 70% and 20%, respectively.

Figure 4(A) illustrates the intervention levels in each cluster when Abate is at the highest level (50%) and as the upper bound on tethering increases. Similarly, Figure 4(B) reports the optimized Abate levels across clusters as the upperbound on Abate increases and tethering is fixed at 95%. As more flexibility is allowed in intervention levels, more tethering resources are devoted to the Central and West clusters and more Abate resources are devoted to the West and East clusters. This is also the case in the optimized solution, shown in Figure 5, where the total percentage of infected dogs after five years of intervention is 2.07%, which translates to 220 fewer dogs infected compared to that under the uniform strategy.

**Figure 4.**
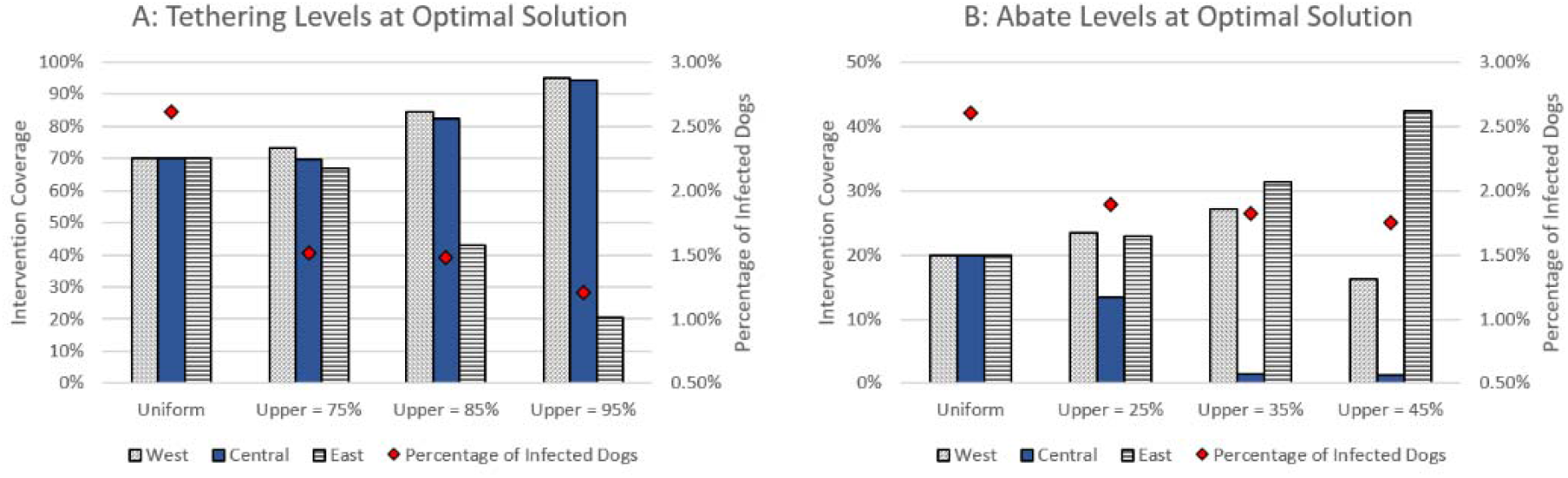
(A) Tethering levels in all clusters under different intervention upper bounds with Abate fixed at 50%; (B) Abate levels in all clusters under different upper bounds with tethering fixed at 95%.

**Figure 5.**
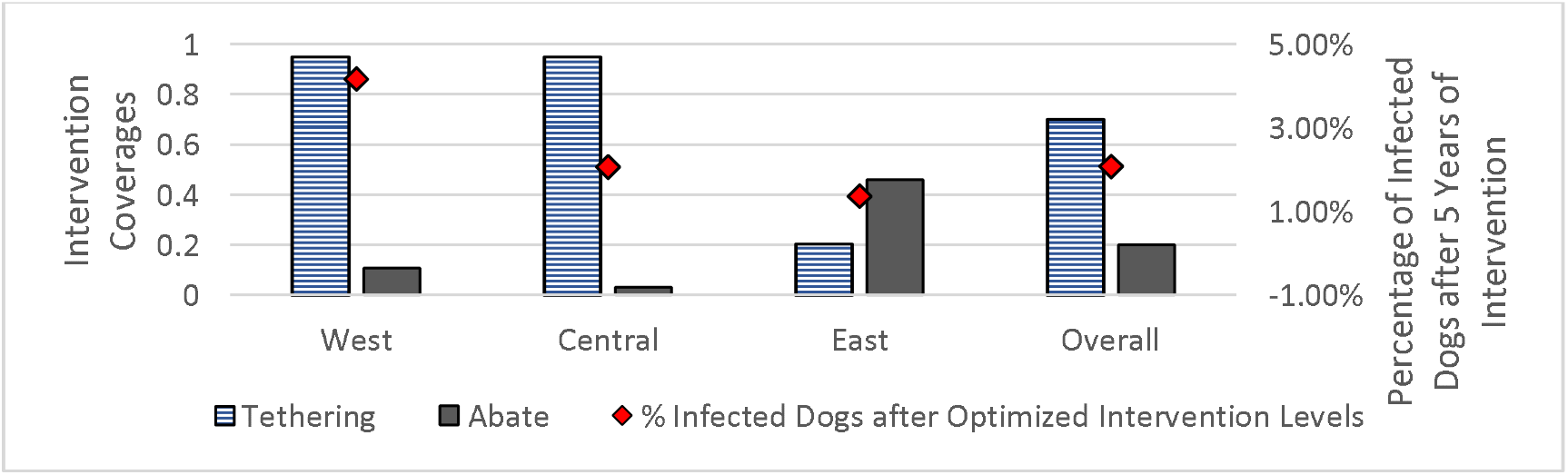
Optimized strategy with average resource levels of 70% and 20% for tethering and Abate, respectively.

For the optimized strategy, the percentages of dogs infected in the West, Central, and East, respectively, are:

- 4.17%, 2.07%, and 1.37% with average (across clusters) tethering and Abate levels of 70% and 20%, respectively.
- 2.81%, 2.07%, and 1.37%. with average tethering and Abate levels of 80% and 30%, respectively. Details are included in Supplemental Materials Section 7.

Sensitivity analysis is included in the Supplemental Materials Section 8. Increasing Abate levels in any cluster above that assigned in the optimized solution would reduce infections, but most notably in the West cluster. Increasing tethering levels in the East cluster beyond the optimized value of 20% would have limited impact on dog infections.

#### Optimized Strategies considering Allocation and Outcome Fairness

To consider allocation fairness, we assigned penalties to tethering level differences among clusters; optimized cluster-specific intervention strategies for each penalty are reported in Table 4. As the penalty increases from 0 to 20, the *allocation fairness* increases (i.e., tethering levels become similar across clusters); however, the percentage of infected dogs increases from 2.07% to 2.61% after five years of intervention. In addition, as the penalty increases, the range in the percentage of infected dogs across the clusters increases slightly (from [1.37%, 4.31%], or 2.9 percentage points difference between clusters, to [2.00%, 5.12%], or 3.1 percentage points difference between clusters). Higher allocation fairness leads to lower outcome fairness in this case. Note also that the outcomes for the Central cluster are worse for each case compared to uniform intervention levels.

The optimized strategies when considering *outcome fairness* (similarity or equality of outcomes between clusters) is displayed in Figure 6. In this approach, no cluster is allowed to have worse outcomes than under the uniform strategy. By prioritizing outcome fairness, the decrease in the percentage of dogs infected across the clusters goes from 5.12%, 2.09%, and 2.00% in the uniform strategy to 4.40%, 2.09%, and 1.84% for the West, Central, and East clusters, respectively. The overall percentage of infected dogs decreases from 2.61% (uniform) to 2.32%.

**Figure 6.**
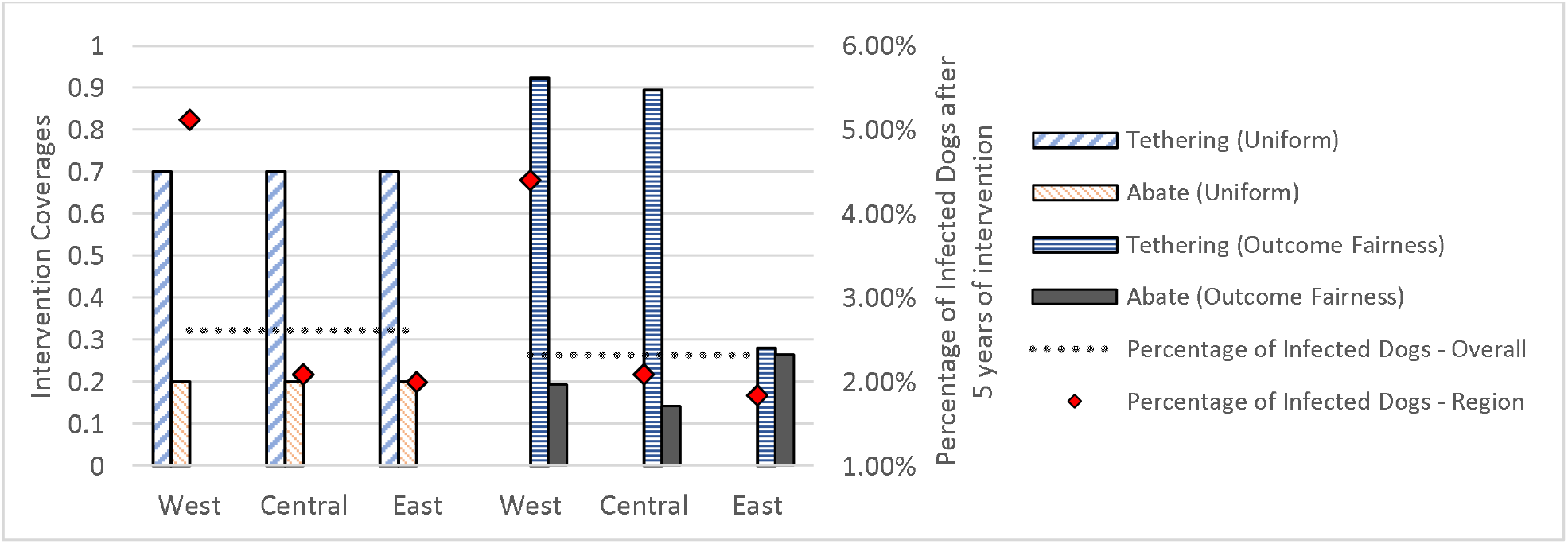
Optimized strategies by cluster under the uniform strategy and with constraints to maximize outcome fairness.

## DISCUSSION

Tethering and Abate are important interventions for reducing GW infections in dogs in Chad. We found that high levels of interventions are needed to reach elimination. When resource limitations prevent high intervention levels in all clusters, using an optimization-based strategy which accounts for heterogeneity across clusters can reduce infections faster than treating clusters equally. Data-driven analytics-based decision making can be a significant aid in the effort to achieve elimination.

If interventions are implemented uniformly across clusters, the results suggest that a minimum of 70% tethering and 35% Abate are needed to reduce infections over time. As shown in Table 2, when tethering or Abate drop below 70% or 35%, respectively, there is a large marginal increase in the number of infected dogs. We found that Abate is more effective than tethering at the same coverage level: for example, under the uniform strategy, 55% tethering combined with 75% Abate results in 334 infected dogs whereas 75% tethering combined with 55% Abate results in 611 infected dogs. In recent years, Chad’s GWEP has been able to increase the number of water sources treated and villages reached, from monthly Abate treatments in 83 villages in 2018 to 600-660 villages in 2021;^22^ at least one water source was treated in 61% of villages with infections by December 2021.^23^

More dog infection cases can be prevented if interventions are targeted using optimization methods rather than uniformly. In the optimized solution, only 2.07%, or 911 dogs, would be infected after five years of intervention, compared to 2.61%, or 1,148 dogs, if interventions are uniform across clusters. This strategy provides a similar level of infection control as 70% uniform tethering and 35% uniform Abate (Table 2). In the optimized solution, higher levels of tethering are needed in the West and Central clusters, but low Abate levels, and then high Abate and lower tethering in the East cluster. The Central cluster connects with each of the other clusters and thus has the highest number of dogs going in and out of the cluster. Under historical intervention levels, we found it is more effective to use a very high tethering level in the Central cluster and minimal Abate. The specific intervention breakdown depends on the capacity for implementing interventions.

Although the optimized strategy achieves the best outcome overall, fairness could be another important consideration. We demonstrated several approaches to fairness. *Allocation fairness* seeks to minimize the differences in intervention levels across clusters to address potential perceptions of favoritism. However, intervention levels which are very similar across clusters are associated with increases in the number of infections overall. Treating all clusters equally will not lead to elimination as quickly as targeting interventions where they will make the most impact. With higher tethering levels in the West and Central clusters and higher levels of Abate in the East cluster, we can maintain *outcome fairness* (Figure 6) where the outcome for each cluster is at least as good as it would be under a uniform (or equal) strategy.

### Limitations

We used data on past interventions, which could be incomplete or inaccurate if compliance with tethering was different than estimated. However, we also considered the cases where tethering is at extreme upper and lowerbound levels. The simulation models assume that tethered dogs are kept on a leash 100% of the time and prevented from exuding worms into water sources. Data about tethering behavior is limited. In reality, tethered dogs may be released for part of the day/night and increase the infectivity of the water sources when exuding worms. We also assume that tethered dogs will be fed clean (uninfected) food and water; however, infections can occur if dogs are fed uncooked aquatic animals.^17,24^ Other animals may exude worms into water sources and increase water infectivity. We assume that no humans will exude worms into the water sources based on extensive efforts in country to educate communities about guinea worm disease and how to reduce infections.

Interventions for GW elimination are important and impactful but overall insufficient if continued at historical levels; even an optimized strategy achieving these levels (on average, across different clusters) is likely to take many years to reach elimination within Chad. This work suggests that a targeted approach can be helpful towards reaching elimination faster. The proposed methods enable decision-makers to identify optimized stategies and assess the trade-off between allocation and outcome fairness. As suggested by Perini et al. ^7^, the model can be generalized to almost any population of definitive hosts with modest adjustments to the parameters and environmental factor distribution. Strategies for determining intervention levels provided in this paper can also be applied in the infection control of other hosts. Future work includes the development of a decision-support tool for decision makers and GWEP personnel in Chad to use as an aid for evaluating various targeted strategies on forecasted dog infections.

## Data Availability

All data produced in the present study are available upon reasonable request to the authors.

## Acknowledgments

This study was supported by a grant from the Carter Center. This research was also supported in part by the following Georgia Tech benefactors: William W. George, Andrea Laliberte, Joseph C. Mello, Richard “Rick” E. and Charlene Zalesky. Dr. Pinar Keskinocak is the William W. George Chair and Professor in the H. Milton Stewart School of Industrial and Systems Engineering at Georgia Institute of Technology. Dr. Julie Swann is the department head and A. Doug Allison Distinguished Professor of the Fitts Department of Industrial and Systems Engineering at North Carolina State University. Dr. Keskinocak and Dr. Swann are the co-founders of the Center for Health and Humanitarian Systems, one of the first interdisciplinary research centers on the Georgia Tech campus.

## Notes

### Competing Interest Statement

The authors have declared no competing interest.

### Funding Statement

This study did not receive any funding.

