## Supplemental Materials for "Evaluating the Effectiveness of Potential Interventions for Guinea Worm Disease in Dogs in Chad Using Simulations"

**Appendix**

#### Section 1: Regional Clustering and Parameter Calibration

The Chad GW Eradication Program (GWEP) documented GW infections within 5 regions, 19 districts, 88 zones, and 1674 villages of Chad. We considered "district" as the smallest units for clustering to balance the level of complexity and data quality (of GW infection in each unit). We used the relative worm emergence per month, latitude, longitude, position along the Chari river and elevation of each district center as inputs for K-means to generate clustering results. Given a point in space, the nearest point on the river is found by computing the intersection between a linear approximation of the river and the perpendicular line including the point. Notably, the timing of worm emergence carries 3 times the weight of the geographic data, cumulatively. We compare the results from K-means (k = 4) with the 5 administrative regions in Chad (Figure S1). Since the distances from SLM region to any other regions, and to the river, is more than 40 km, we consider the districts within the SLM region to be isolated from the others.

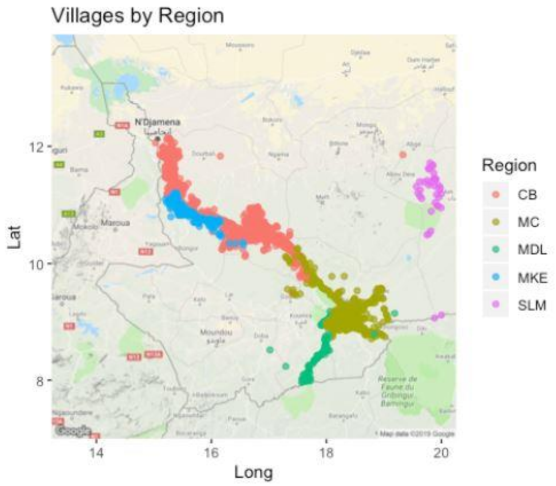


*Figure S1 Villages by administrative regions in Chad.*

The number of worms exuded from 2016 to 2018 within each cluster are plotted in Figure S2.

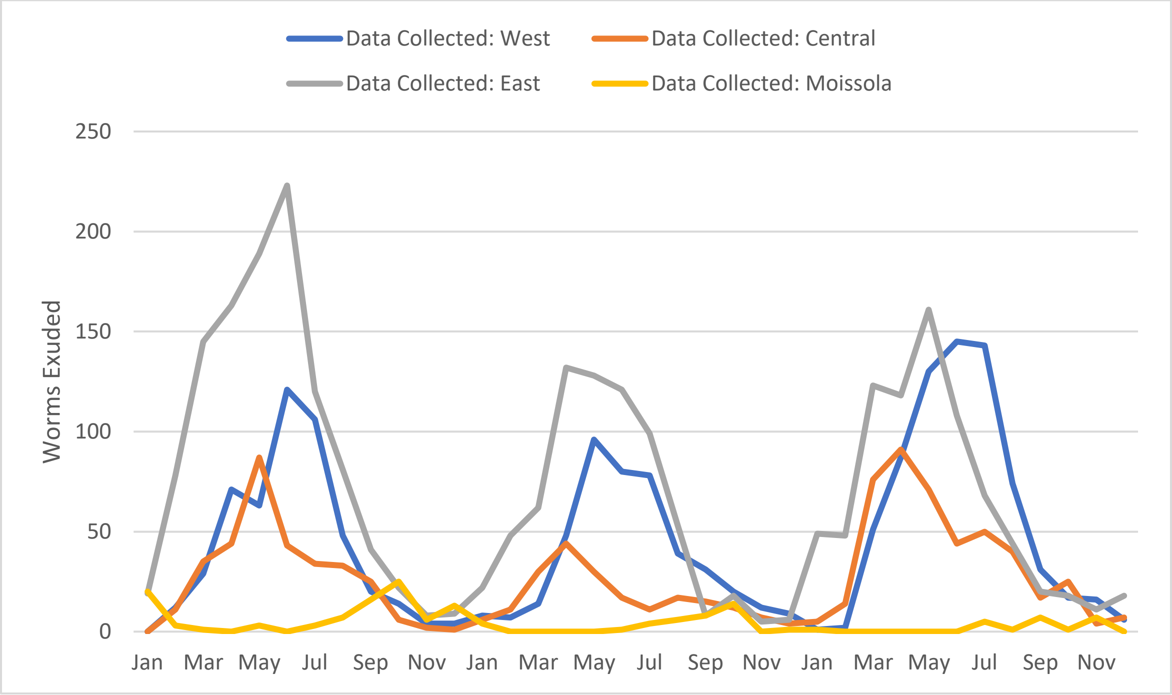


*Figure S2 Regional dog infection data for years 2016-2018*

The Moissala Cluster differs from the other three in both infection seasonality and infection magnitude. Additionally, there are 13 months within the three-year-period where no exuding dogs are documented in this cluster. The Moissala cluster is too small to be considered as an individual cluster and its model is hard to validate due to insufficient data. Thus, we removed this cluster from our model.

In order to calibrate parameters for each cluster, we first integrate the number of worms emerging from dogs per month and the number of dogs with an emerging worm per month for the years 2016-2018 for each cluster (Figure S2 and Figure 3). Then, we collected the percentage of tethering conducted for each region per year (Table S1). Note that, starting from 2017, tethering status for a certain number of infected dogs is not documented, thus we calculated ranges for the actual tethering coverage.

*Table S1 Tethering coverage for each cluster in Chad*

| Cluster | Tethering Coverage | | |
|---|---|---|---|
| | **2016** | **2017** | **2018** |
| West Chari | 0.63 | [0.43, 0.83] | [0.43, 0.88] |
| Central Chari | 0.74 | [0.62, 0.90] | [0.49, 0.88] |
| East Chari | 0.72 | [0.40, 0.86] | [0.32, 0.95] |

Even with the best set of parameters reported in Perini et al. [7], the simulation model failed to capture the difference in GW infection cases between clusters. Thus, for each cluster, we calibrate parameters for the infectivity curve separately. We use a sigmoidal function to represent the probability that a dog is infected on a particular day after interacting with a water source; the basic sigmoidal function, when there are n infective copepods in the water source (i.e., worm burden), is modeled as $f(n) = \frac{1}{1+e^{-C(n-D)}}$, where C and D represent the curvature parameter and inflection point, respectively. The basic infectivity function (i.e., the infectivity of the water source given n) is $F(n) = [f(n) - f(0)](F_U - F_L) + F_L$ , where $F_L$ and $F_U$ represent the minimum and maximum rates of infection, respectively. Seasonality is incorporated in the final infectivity function $F_{inf}(n, m, y) = w_m \times (1 - a_y) \times F(n)$ , where $w_m \in [0,1]$ is the environmental factor in month m, and $a_y \in [0,1]$ represents the intervention coverage of Abate in year y. We incorporate environmental factors developed by Perini et al.[7] (see Table S2) which are based on daily precipitation and temperature measurements from all 17 weather stations in Chad provided through the National Centers for Environmental Information's Climate Data Online database [20].

*Table S2 Environmental factors*

| Month | Jan | Feb | Mar | Apr | May | Jun | Jul | Aug | Sep | Oct | Nov | Dec |
|---|---|---|---|---|---|---|---|---|---|---|---|---|
| **Factor** | 0.0981 | 0.2481 | 0.2069 | 0.9669 | 0.9984 | 1.00 | 0.4977 | 0.5968 | 0.3446 | 0.1808 | 0.2520 | 0.1018 |

Empirical data is more likely to be underreported than overreported for both the number of exuding dogs and GWs. Thus, we calibrate the model parameters by minimizing the combined weighted mean squared error (MSE) of the exuding dogs and worms (simulated and observed), with underestimation weighted more heavily than overestimation (4 to 1).

We created three scenarios for each cluster, where the tethering coverage of 2017 is the minimum, mean and the maximum of the documented range. For each scenario, we calibrate parameters for infectivity curves. The simulation is initialized with data from 2016 (number of infected dogs and number of emerging worms), and the remaining data from 2017-2018 are used for calibration. We take 20 replications for each simulation scenario. Parameter searches are performed using the cross-entropy method; for each iteration, we sample 50 potential solutions, and for each sample we simulate twenty replications. We stop when we reach 50 iterations or when the maximum difference between two iterations is very small.

The infectivity curves for each cluster and each tethering case (minimum of range, mean, and maximum) are shown in Figure S3, and simulation results for each cluster and tethering option are shown in Figure S4. The figure reports the MSE for dogs and worms for each case. Because of the reduced MSE for worms and dogs in the Central cluster for tethering equal to the mean of the documented ranges, and the comparable MSE for dogs in the East and West Clusters across the three tethering options, and for consistency across clusters, we chose the mean tethering case for our analysis. The final parameters for the infectivity curves chosen for each cluster (mean tethering) are shown in Table S3.

*Table S3 Infectivity parameters for each cluster.*

| Cluster | Minimum Infectivity Rate | Maximum Infectivity Rate | Inflection Point | Curvature |
|---|---|---|---|---|
| **East** | 1.53e-04 | 2.472e-03 | 125 | 0.1661 |
| **Central** | 1.55e-04 | 3.983e-03 | 88 | 0.1210 |
| **West** | 3.4e-04 | 4.265e-03 | 78 | 0.0668 |

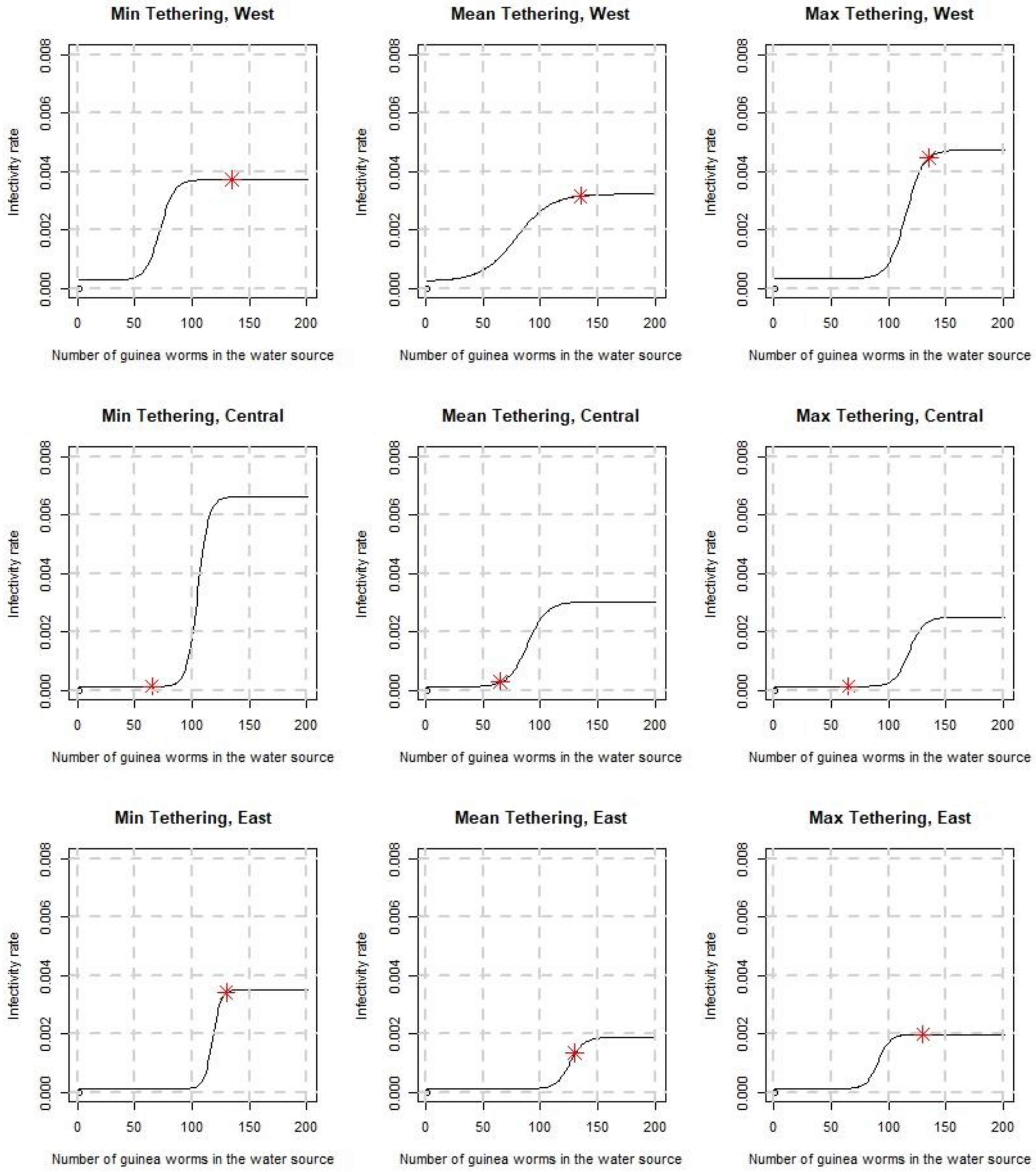


*Figure S3 Infectivity curves for each region and tethering level (min, mean, and max of documented range), considering abate coverage in 2018. The star corresponds to a status quo of the number of GWs in the fourth year (calculated as the maximum worms exuded across all months in 2019).*

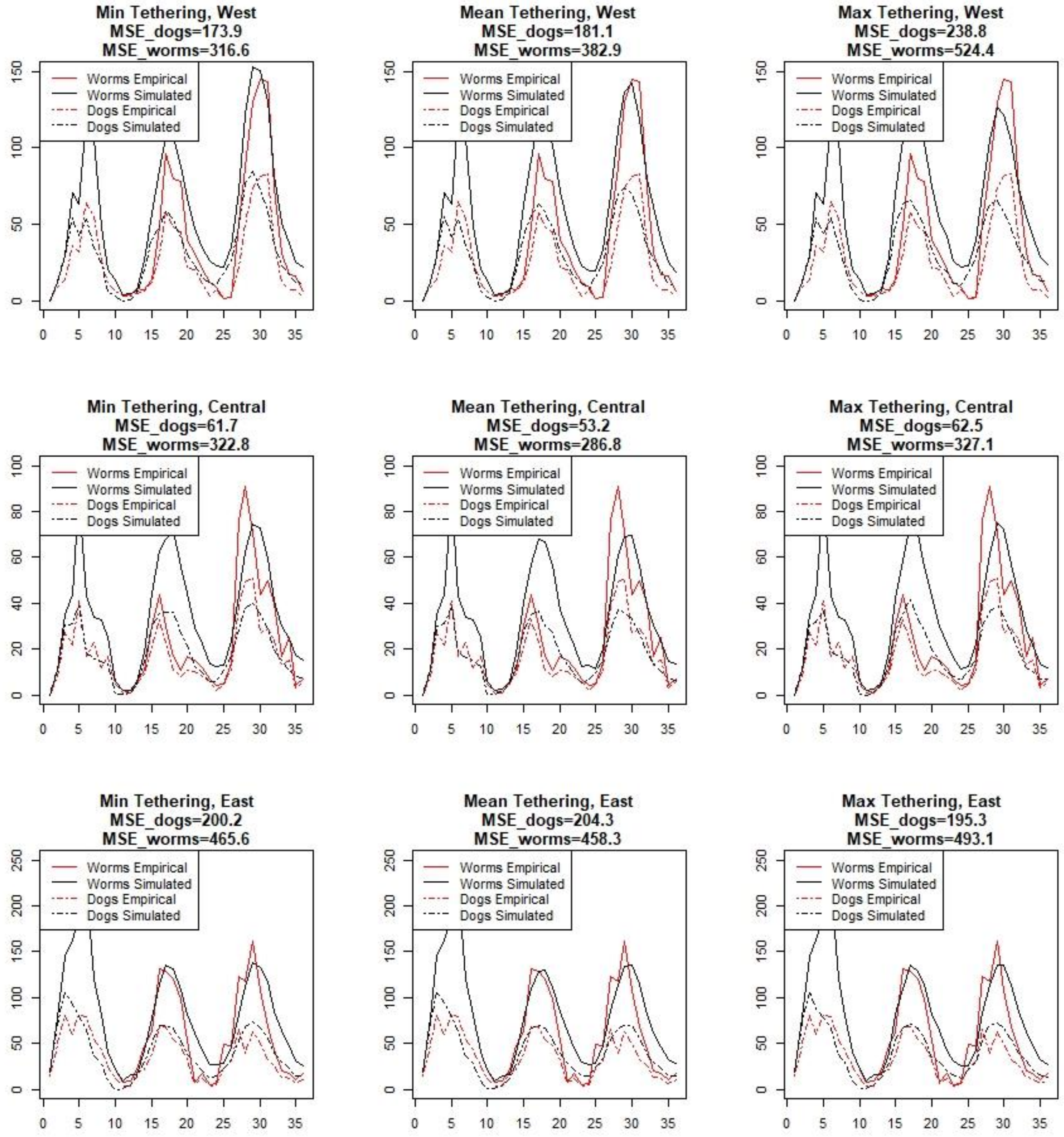


*Figure S4 GWs exuded and dogs infected (empirical and simulated) for the West, Central, and East clusters, under different tethering options (min, mean, and max of documented range).*

#### Section 2: Simulation Model

**Algorithm 1** Pseudo code for the simulation

---

nbDogs = [1:N]

*randvec* = vector of uniformly distributed random values in (0,1) of length N

**Set** initial number of dogs infected, worms exuded per region based on empirical data

**Define** water contact probability DW(i,r) for each dog i and water source r (region) based on travel behavior

**for** iteration from 1 to 20

  *#Initialize dog infections based on empirical data:*

  **Choose** random sample of dogs per region to be infected

  **Assign** worms per dog based on worms per dog distribution and **calculate** day each worm will exude

  **for** d from 1 to 360*(simulation years)

    *#Update number of worms in the water sources:*

    **Define (**per region r) the number of mature worms in the water source =

          previous number of worms – worms that die on day d + worms reaching maturity on day d

    For each region r:

      **Identify** any dogs from r exuding worms on day d

        **Update** number of worms exuded and number of dogs exuding

        For each identified dog exuding worms on day d:

          **Assume** dog is tethered (for 30 days) if random value in (0,1) ≤ tethering probability of r in current year

    *#Determine the number of new Guinea worms in the water on day d:*

    For each region r:

      **if** the current month is in the dry season, the number of new Guinea worms in the water source

        = the number of exuding *non-tethered* dogs with water contact probability with region r greater than 10%

      **else** = the number of exuding *non-tethered* dogs from region r only

**end if**

**Update** lists of expected worm maturity and death days

**Determine** ABATE impact:

**Define** number of Guinea worms killed k = random number from binomial distribution (n = number of Guinea worms in the water in r, p = abate level in r in the current year).

**Update** number of Guinea worms in the water in r, removing k randomly chosen matured Guinea worms from the water

*#Calculate infectivity of the water sources:*

For each region r:

**Calculate** WInf(r) **=** water infectivity rate for region r using environmental factor, abate level, infectivity parameters, and the number of Guinea worms in the water

*#Determine the dogs from r that are infected by a water source on day d*

**Define** a threshold value V for each dog i from r which accounts for contact probabilities and water infectivity:

**If** the current month is in the dry season, then dog i may have contact with multiple water sources (regions):

V = DW(i,1)* WInf (1) + DW(i,2)* WInf (2) + DW(i,3)* WInf (3)

**otherwise,** V = DW(i,r)* WInf (r)

**Assume** dog i is infected on day d if randvec(i) < V (*ignore tethered dogs*)

**Assign** worms per dog based on worms per dog distribution, and calculate exuding days

**Update** lists of infected dogs and exuding days

**end for**

**end for**

**return** average worms exuded and dogs exuding by region and day

**Section 3: Optimized Intervention Strategies**

We explored four methods for determining resource levels for clusters, assuming resource capacities enabling 70% tethering and 20% Abate levels across all clusters combined:

1. We used the CE method to search for the optimized intervention strategy for each individual cluster under cluster-specific levels of 70% for tethering and 20% for Abate.
2. We then used the CE method to determine the optimized intervention levels under cluster-specific levels ranging from 75% to 95% for tethering and 25% to 50% for abate. Lower upperbounds on these levels resulted in improved equality of tethering and Abate levels between clusters, or *allocation fairness.*
3. To improve allocation fairness (method 2), we added penalties (ranging from 0 to 20) in the model objective function:
    - As the penalty increases from 0 to 20, the model attempts to achieve similar intervention levels across clusters, unless the benefit from different levels between clusters outweighs the penalty.

   We maintained cluster-specific upper bounds (95% and 50% for tethering and Abate, respectively). We searched for optimized intervention strategies using the CE method under each penalty weight.
4. To improve *outcome fairness* of targeted levels of interventions, we added constraints to the model to ensure that all clusters achieve a lower percentage of infections after five years of intervention compared to the percentage of infections under a uniform strategy. We used the CE method to search for the optimized intervention strategy.

We repeated the first two methods with increased intervention resource levels (80% tethering and 30% Abate across all clusters combined).

For each scenario, multiple iterations are needed. For each iteration, we sampled 200 potential solutions, and for each sample we simulated twenty replications of five years of intervention. The CE method ranked the sample solutions by performance to inform the choices of the next potential solutions. We stopped after reaching 20 iterations or when the maximum difference between two iterations was very small.

#### Section 4: Optimization Model

*Table S4 Notation for the simulation optimization model for determining the best alternate strategies which are at least as good as the uniform strategy.*

| **Notation** | **Description** |
| --- | --- |
| Decision Variables | |
| $T_w \in [0,1]$ | Tethering coverage in the West cluster. |
| $T_c \in [0,1]$ | Tethering coverage in the Central cluster. |
| $T_e \in [0,1]$ | Tethering coverage in the East cluster. |
| $A_w \in [0,1]$ | Abate coverage in the West cluster. |
| $A_c \in [0,1]$ | Abate coverage in the Central cluster. |
| $A_e \in [0,1]$ | Abate coverage in the East cluster. |
| Auxiliary Variables | |
| $\mathrm{Inf}_i(\vec{T}, \vec{A})$ | Total number of dog infections in cluster i, with intervention strategy $\vec{\boldsymbol{T}}, \vec{\boldsymbol{A}}$ |
| $\vec{T}$ | Tethering strategy: $\vec{\boldsymbol{T}} = [\boldsymbol{T_w}, \boldsymbol{T_c}, \boldsymbol{T_e}]$ |
| $\vec{A}$ | Abate strategy: $\vec{\boldsymbol{A}} = [\boldsymbol{A_w}, \boldsymbol{A_c}, \boldsymbol{A_e}]$ |

| | |
|---|---|
| $\overrightarrow{T_U}$ | Uniform Tethering strategy |
| $\vec{A}_{\mathrm{U}}$ | Uniform Abate strategy |
| $P_i$ | Dog population in cluster i |
| $T_u, A_u$ | Upper bound for tethering/Abate levels |
| $\mathrm{C_T}, \mathrm{C_A}$ | Resource capacity for tethering/Abate |
| $M$ | Penalty weight [2,4,6,8,10] |
| $D$ | Maximum tethering difference across clusters: Max $[\boldsymbol{T_w}, \boldsymbol{T_c}, \boldsymbol{T_e}]$– Min $[\boldsymbol{T_w}, \boldsymbol{T_c}, \boldsymbol{T_e}]$ |

$$\mathbf{Min}\ \frac{\sum_{i\in\{W,C,E\}} Inf_i(\vec{T},\vec{A})}{\sum_{i\in\{W,C,E\}} P_i} + [\left(\frac{M}{1000}\right) * D]$$

$$\mathbf{s.t.}\ \ \frac{\mathrm{T_w} + T_c + T_e}{3} = C_T$$

$$\frac{\mathrm{A_w} + A_c + A_e}{3} = C_A$$

$$\frac{Inf_i(\vec{T},\vec{A})}{P_i} \le \frac{Inf_i(\overrightarrow{T_U},\vec{A}_U)}{P_i}$$

$$\mathrm{T_w}, \mathrm{T_c}, \mathrm{T_e} \le T_u$$

$$\mathrm{A_w}, \mathrm{A_c}, \mathrm{A_e} \le A_u$$

**Section 5: Traveling Behavior of Dogs**

Research on the ecology of Guinea Worm infection in dogs suggests that 80% of dogs visit ponds that are within a 100-meter-range from the dog owner's house [17]. Additionally, a dog's mean travel range is 4.4 square kilometers in Chad, much higher in dry season than in wet season [17]. Based on the geographical data, we assigned groups to dogs within each cluster. The interactions between clusters are:

1. Dogs do not use the water source from the other clusters during the wet season (from June to October).
2. Dog group I (40%) in the West Cluster uses the Central water source as their alternative with a probability of 20% during the dry season; group II (60%) uses the West water source with 100% probability.
3. Dog group I (10%) in the East Cluster uses the Central water source as their alternative with a probability of 20% during the dry season; group II (90%) uses the East water source with 100% probability.
4. Dog group I (40%) in the Central Cluster uses the West water source as their alternative with a probability of 20% during the dry season. Dog group III (10%) uses the East water source as their alternative with a probability of 20% during the dry season. Dog group II (50%) uses the Central water source with 100% probability.
5. Dogs do not switch groups.

#### Section 6: Trends Over Time

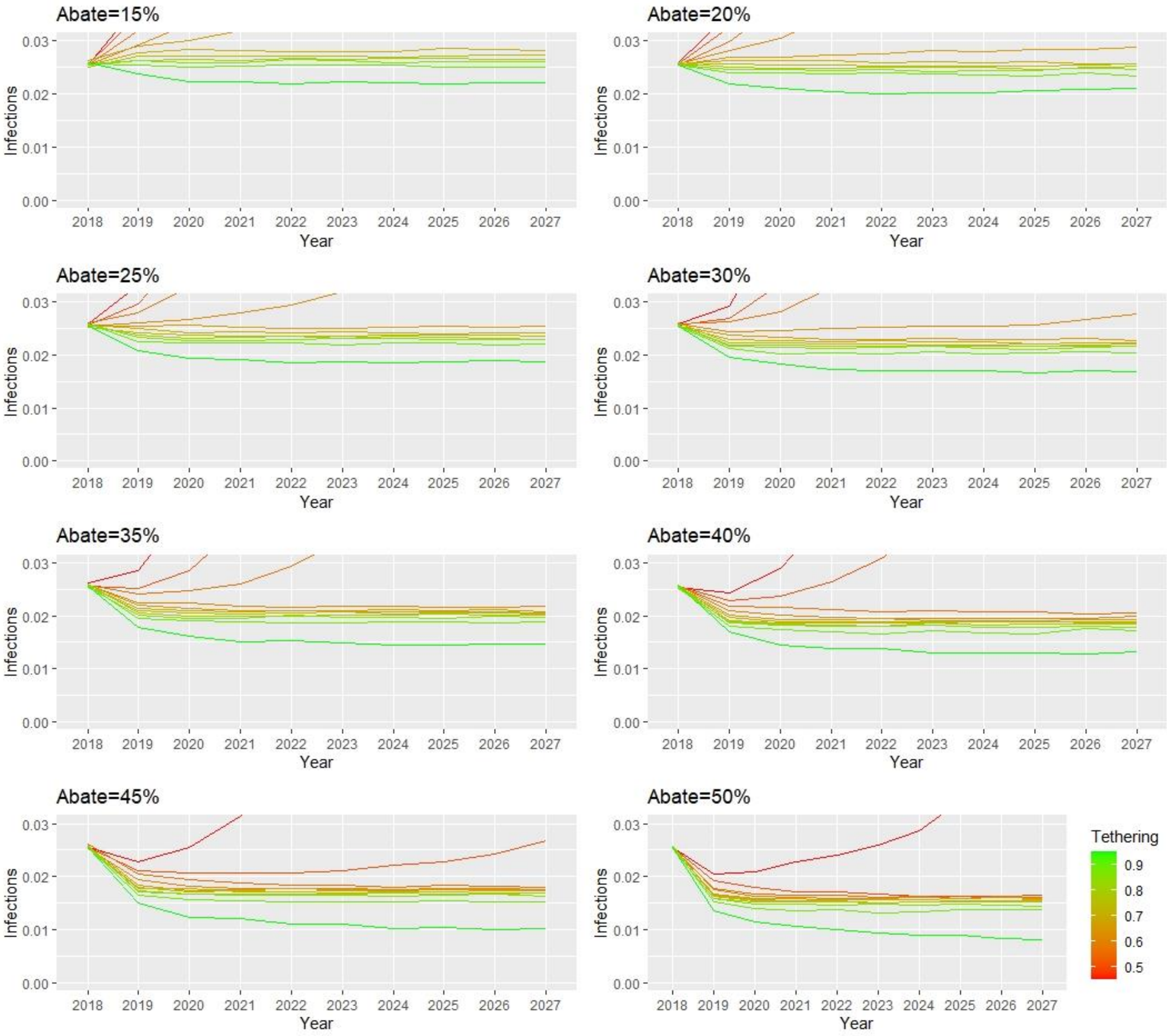


*Figure S5 The percentage of dog infections over time and across all clusters for varying levels of abate treatment (15-50%) and tethering.*

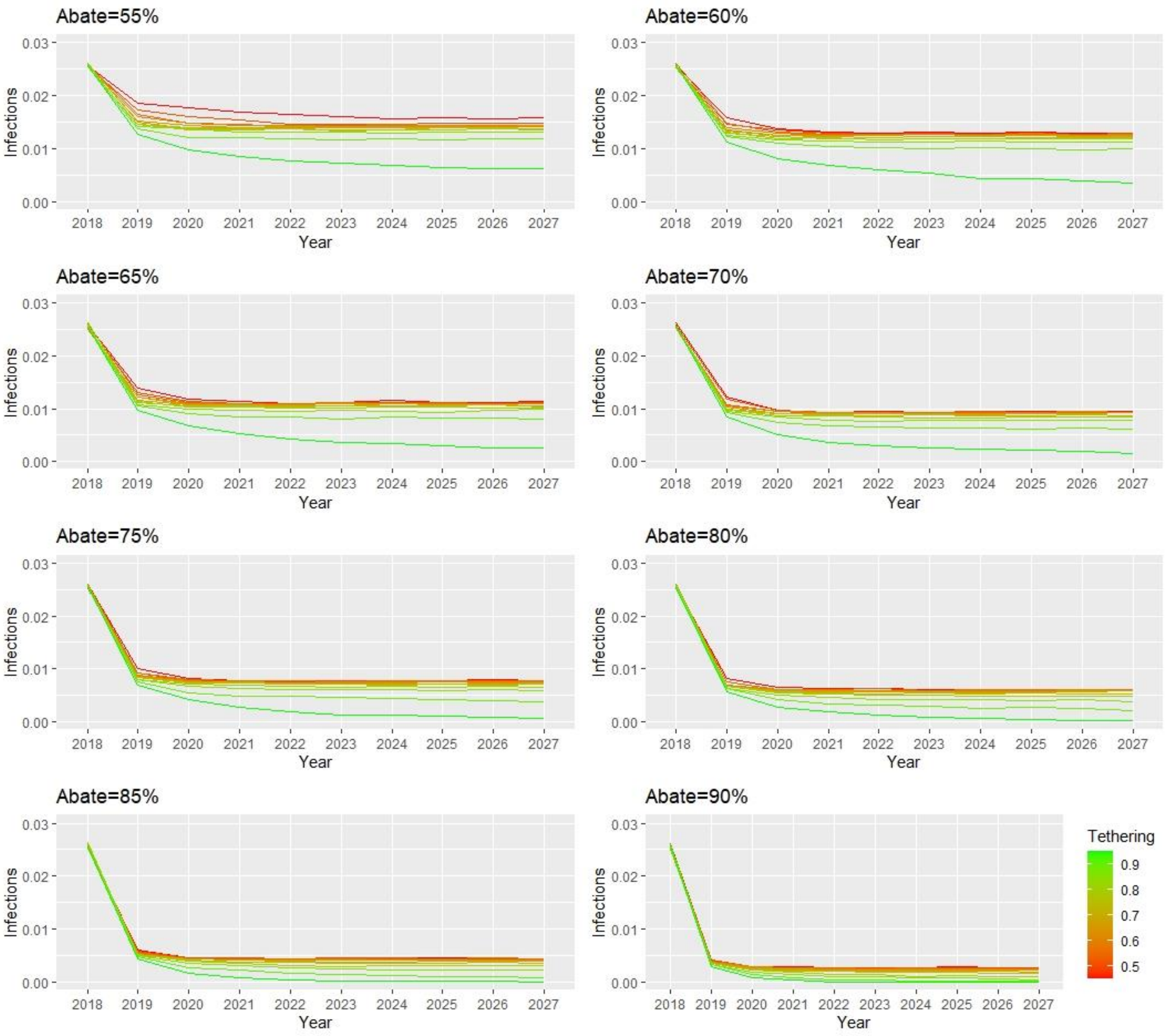


*Figure S6 The percentage of dog infections over time and across all clusters for varying levels of abate treatment (55-90%) and tethering.*

#### Section 7: Regional Strategy with Increased Resources

We determined optimized intervention strategies under varied cluster-specific intervention upper bounds and increased overall resources at the national level (80% national coverage for tethering and 30% national coverage for Abate). Optimized strategies under various intervention upper bounds are shown in Table S5. The optimized strategy without cluster-specific upper bounds is shown in Figure S7.

*Table S5 Percentage of dog infections after five years of the optimized intervention strategy under different cluster-specific intervention upper bounds, with national upper bounds of 80% for tethering and 30% for Abate.*

| **Resource Level:** | | **Upper Bound of Abate** | | | | |
|---|---|---|---|---|---|---|
| [0.80,0.30] | | 0.30 | 0.35 | 0.40 | 0.45 | 0.50 |
| **Upper Bound of Tethering** | 0.8 | 2.14% | | | | |
| | 0.85 | | 2.14% | 2.14% | 2.08% | 2.08% |
| | 0.9 | | 2.14% | 2.01% | 1.98% | 1.88% |
| | 0.95 | | 2.10% | 1.80% | 1.75% | 1.70% |

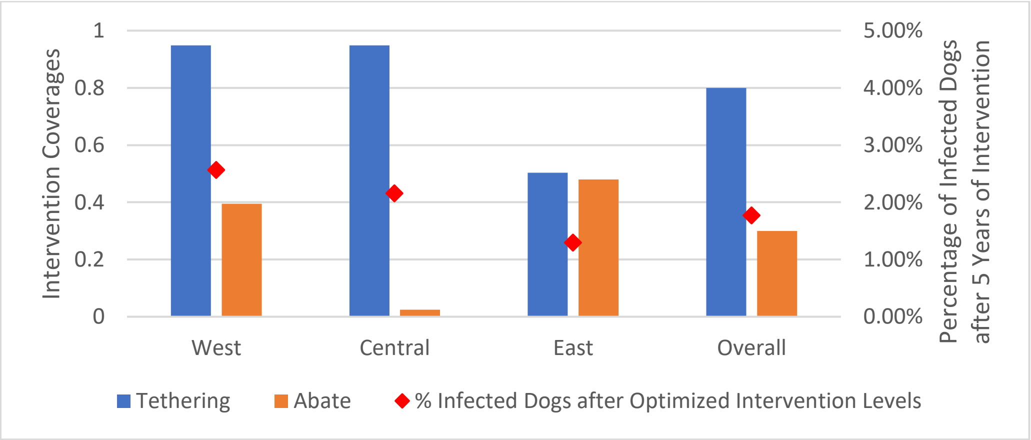


*Figure S7 Optimized intervention strategy under a national resource capacity of 80% for tethering and 30% for Abate*

#### Section 8: Sensitivity Analysis on Intervention Coverages

We select the optimized intervention strategy as a baseline for this set of sensitivity analyses. We vary each cluster-specific intervention coverage from 0% to 96% while the other intervention levels remain the same. The results are seen in Figure S8 and Figure S9. We observe in Figure S8 that in each cluster, when all levels are set to that of the optimized strategy, no additional tethering in an individual cluster will have a significant effect on infection control. However, with other optimized values fixed, the tethering levels in the Central cluster can be reduced to about 60% and achieve the same results. In the East cluster, when tethering is set to the optimized level of about 20%, we see in Figure S9 that Abate resources below about 40% greatly increase infections. There are marginal reductions in infections which are mostly linear when Abate increases in each cluster.

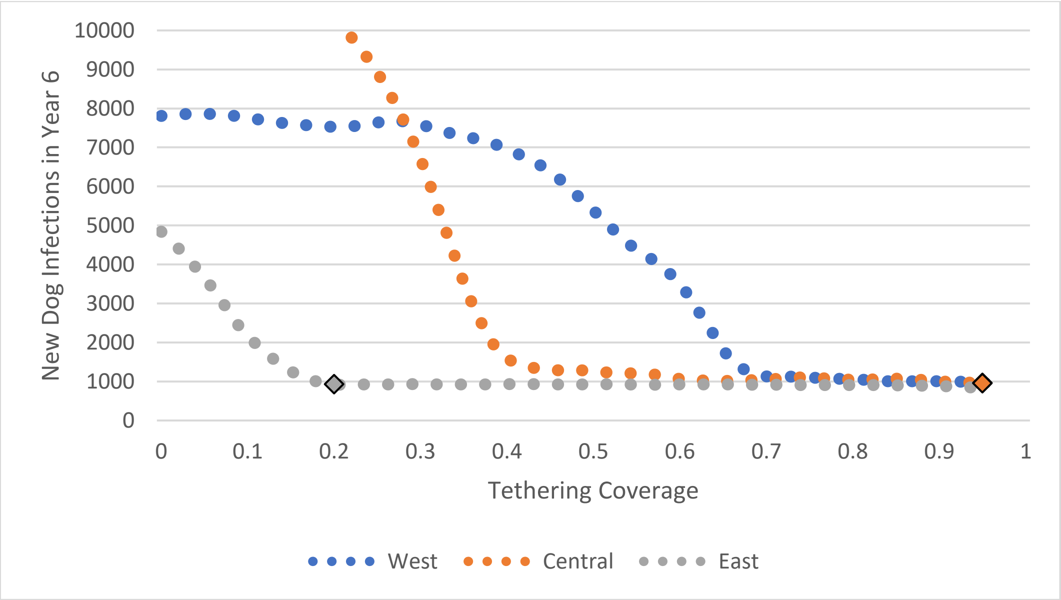


*Figure S8 Sensitivity analysis on tethering coverage and corresponding new infections after five years of intervention in the West, Central and East clusters. The diamonds indicate the optimized tethering level per cluster. Abate coverage levels are set to the optimized intervention strategy levels.*

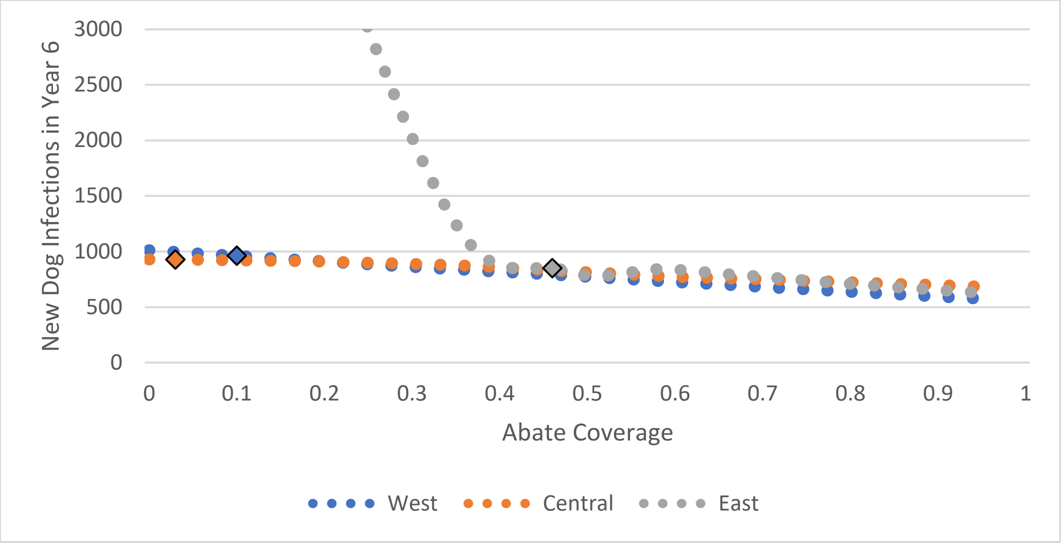


*Figure S9 Sensitivity analysis on Abate coverage and corresponding new infections after five years of intervention in the West, Central and East clusters. The diamonds indicate the optimized Abate level per cluster. Tethering coverage levels are set to the optimized intervention strategy levels*

### Section 9: Alternate Calibrations

*Table S6 Percentage of dogs infected after uniform interventions for 5 years, with parameters calibrated based on the* ***tethering range lowerbound****.*

| | | Abate Levels (Uniform) | | | | | | | | | | | | | | | | |
|---|---|---|---|---|---|---|---|---|---|---|---|---|---|---|---|---|---|---|
| | | 15% | **20%** | 25% | 30% | 35% | 40% | 45% | 50% | 55% | 60% | 65% | 70% | 75% | 80% | 85% | 90% | 95% |
| **Tethering Levels (Uniform)** | 45% | 44.98% | 38.75% | 39.13% | 28.78% | 26.43% | 16.74% | 13.77% | 6.79% | 7.70% | 5.79% | 2.09% | 1.61% | 0.78% | 0.63% | 0.45% | 0.28% | 0.11% |
| | 50% | 37.06% | 33.55% | 29.39% | 20.33% | 18.63% | 12.03% | 10.98% | 6.61% | 5.03% | 4.17% | 2.67% | 2.00% | 0.79% | 0.64% | 0.46% | 0.28% | 0.10% |
| | 55% | 27.15% | 22.22% | 17.60% | 15.69% | 10.85% | 8.19% | 5.10% | 4.69% | 2.28% | 2.20% | 1.19% | 0.94% | 0.79% | 0.62% | 0.45% | 0.27% | 0.09% |
| | 60% | 12.70% | 13.23% | 13.47% | 9.23% | 6.92% | 5.38% | 4.53% | 3.33% | 1.93% | 1.44% | 1.11% | 1.05% | 0.78% | 0.61% | 0.45% | 0.26% | 0.08% |
| | 65% | 11.66% | 7.36% | 5.59% | 4.08% | 4.44% | 3.60% | 3.85% | 2.04% | 1.75% | 1.28% | 1.12% | 0.95% | 0.77% | 0.59% | 0.42% | 0.25% | 0.07% |
| | **70%** | 5.83% | 4.63% | 4.25% | 3.64% | 2.85% | 2.80% | 2.38% | 1.60% | 1.44% | 1.26% | 1.08% | 0.94% | 0.77% | 0.58% | 0.40% | 0.22% | 0.06% |
| | 75% | 4.62% | 3.73% | 4.31% | 2.76% | 2.42% | 1.93% | 1.73% | 1.59% | 1.43% | 1.24% | 1.08% | 0.91% | 0.75% | 0.55% | 0.38% | 0.19% | 0.05% |
| | 80% | 3.22% | 3.45% | 2.58% | 2.17% | 2.04% | 1.90% | 1.75% | 1.54% | 1.38% | 1.21% | 1.06% | 0.87% | 0.68% | 0.51% | 0.32% | 0.15% | 0.02% |
| | 85% | 2.63% | 2.53% | 2.36% | 2.18% | 2.01% | 1.86% | 1.67% | 1.50% | 1.35% | 1.17% | 1.01% | 0.82% | 0.62% | 0.42% | 0.25% | 0.10% | 0.01% |
| | 90% | 2.64% | 2.44% | 2.28% | 2.12% | 1.90% | 1.76% | 1.59% | 1.43% | 1.21% | 1.07% | 0.86% | 0.69% | 0.45% | 0.31% | 0.16% | 0.03% | 0.003% |
| | 95% | 2.33% | 2.17% | 1.93% | 1.76% | 1.57% | 1.38% | 1.13% | 0.99% | 0.75% | 0.63% | 0.43% | 0.32% | 0.17% | 0.07% | 0.02% | 0.004% | 0.000% |

*Table S7 Percentage of dogs infected after uniform interventions for 5 years, with parameters calibrated based on the **tethering range upperbound**.*

| | | Abate Levels (Uniform) | | | | | | | | | | | | | | | | |
|---|---|---|---|---|---|---|---|---|---|---|---|---|---|---|---|---|---|---|
| | | 15% | **20%** | 25% | 30% | 35% | 40% | 45% | 50% | 55% | 60% | 65% | 70% | 75% | 80% | 85% | 90% | 95% |
| Tethering Levels (Uniform) | 45% | 14.88% | 8.42% | 3.54% | 2.40% | 2.25% | 2.02% | 1.88% | 1.70% | 1.91% | 1.38% | 1.18% | 1.02% | 0.86% | 0.66% | 0.50% | 0.30% | 0.12% |
| | 50% | 7.00% | 3.31% | 2.52% | 2.37% | 2.17% | 2.02% | 1.84% | 1.69% | 1.56% | 1.35% | 1.17% | 1.01% | 0.84% | 0.67% | 0.47% | 0.31% | 0.10% |
| | 55% | 2.82% | 2.70% | 2.54% | 2.36% | 2.18% | 2.02% | 1.86% | 1.70% | 1.52% | 1.34% | 1.19% | 0.99% | 0.84% | 0.67% | 0.49% | 0.30% | 0.10% |
| | 60% | 2.87% | 2.70% | 2.54% | 2.36% | 2.19% | 2.03% | 1.85% | 1.69% | 1.51% | 1.34% | 1.17% | 1.00% | 0.84% | 0.64% | 0.46% | 0.28% | 0.08% |
| | 65% | 2.86% | 2.70% | 2.53% | 2.32% | 2.21% | 2.03% | 1.84% | 1.69% | 1.53% | 1.34% | 1.18% | 1.00% | 0.83% | 0.64% | 0.46% | 0.26% | 0.07% |
| | **70%** | 2.86% | 2.67% | 2.52% | 2.36% | 2.14% | 2.03% | 1.85% | 1.69% | 1.52% | 1.35% | 1.16% | 0.99% | 0.80% | 0.63% | 0.44% | 0.25% | 0.06% |
| | 75% | 2.87% | 2.72% | 2.48% | 2.35% | 2.15% | 2.04% | 1.83% | 1.68% | 1.47% | 1.33% | 1.15% | 0.98% | 0.80% | 0.61% | 0.42% | 0.21% | 0.05% |
| | 80% | 2.85% | 2.65% | 2.51% | 2.36% | 2.18% | 2.02% | 1.82% | 1.65% | 1.50% | 1.29% | 1.15% | 0.95% | 0.74% | 0.56% | 0.38% | 0.16% | 0.03% |
| | 85% | 2.83% | 2.65% | 2.49% | 2.32% | 2.13% | 2.00% | 1.81% | 1.64% | 1.44% | 1.27% | 1.07% | 0.85% | 0.71% | 0.48% | 0.29% | 0.12% | 0.01% |
| | 90% | 2.80% | 2.61% | 2.44% | 2.24% | 2.11% | 1.91% | 1.71% | 1.57% | 1.32% | 1.14% | 0.96% | 0.72% | 0.60% | 0.36% | 0.19% | 0.07% | 0.003% |
| | 95% | 2.46% | 2.32% | 2.11% | 1.92% | 1.74% | 1.55% | 1.35% | 1.17% | 0.90% | 0.71% | 0.54% | 0.32% | 0.17% | 0.10% | 0.02% | 0.006% | 0.000% |
